# Determinants of obesity among rural adolescents in Vhembe district, Limpopo Province, South Africa

**DOI:** 10.1101/2023.05.12.23289912

**Authors:** Brenda Baloyi, Lindelani Fumudzani Mushaphi, Ngoako Solomon Mabapa

## Abstract

The increased prevalence of obesity is due to a decreased level of physical activity and increased intake of fast food. Furthermore, obesity among children and adolescent is a risk factor for life-threatening conditions including cardiovascular diseases (CVD), Cardio-metabolic disorders, type 2 diabetes mellitus, hypertension, cancer and reproductive disorders. The aim of this study is to describe the determinants of obesity. A cross-sectional study was conducted on a total of 377 adolescents aged 13 to 20 years from 16 secondary schools in Thulamela Municipality, Vhembe District Limpopo Province, South Africa. Information about socio-demographic characteristics, household income, disease family history, and level of education of parents was obtained using a self-administered questionnaire. Anthropometric measurements such as weight, height and waist circumference were taken by trained field workers and body mass index (BMI), and the waist-hip ratio were determined. Biochemical measurements and clinical assessment were done by a professional nurse following standard procedures. The prevalence of obesity is 22.2% in males and 32.6% in females by abdominal obesity by (waist circumference), whilst 11.1% (males) and 28.3% (females) by waist to hip ratio (WHR). Gender (β=0.32, p=0.018, 95%CI); age (β=1.28, p=0.015, 95%CI); source of income (β=3.25, p=0.008, 95%CI) and systolic blood pressure (β=1.04, p=0.01, 95%CI) were associated with obesity. Overweight and obesity were more prevalent in females than in males in Thulamela municipality. There is a need to bring up children and adolescents in a health-promoting environment in an effort to reverse and stop the increasing trend of overweight and obesity.

## INTRODUCTION

Obesity is defined as excessive or abnormal body fat accumulation that may result in serious health problems [1]. Globally obesity among children and adolescents is a serious public health concern [2,3]. Children and adolescents’ obesity indicates a higher risk of transition to adulthood [4-6]. The increased prevalence of obesity is due to decreased level of physical activity and increased intake of fast food [7-9]. Furthermore, obesity among children and adolescent is a risk factor for life-threatening conditions including CVD, Cardio-metabolic disorders, type2 diabetes mellitus, hypertension, cancer and reproductive disorders [10,11].

The prevalence of global obesity has doubled from the year 2000 to 2016, with an estimation of 650 million adults being obese in 2016 [9]. WHO [12] reported that over 340 million children and adolescents were obese in 2016. In 2010, 43 million children were obese with 81% coming from developing countries [13]. Obesity is considered a problem in developed countries however, it is currently on the rise in developing countries [1,14,15]. In South Africa, the prevalence of overweight and obesity has increased from 49.4% in 1980 to 57.8% in 2015 [16]. Studies, [17-20] reported that the prevalence of overweight and obesity is high in girls than in boys. Most studies focused on the prevalence of obesity in urban areas, the ones tracking the determinants of obesity were conducted in urban areas [21]. Little is known about the prevalence and determinants of obesity in the Vhembe district. The aim of this study is to describe the determinants of obesity in Vhembe district, Limpopo Province, SA.

## METHODS

### Participants and study design and sampling

The study is descriptive in nature and followed a cross-sectional design. Slovin’s formula was used to determine the sample size. The formula yielded 400 however 377 adolescents agreed to participate in the study. The participants were selected using stratified random sampling whereby five participants were selected per grade (grades: 8-12). From each secondary school, 25 adolescents were selected. Adolescents attending were free from illness and whose parents gave consent to participate in the study. Pregnant adolescents were excluded from the study. The response rate was 94.25%.

### Study participation

A total of 377 adolescents aged 13 to 20 years from 16 secondary schools participated in the study from February 2018 to August 2018. Participants were recruited in January 2018. The research took place in secondary schools in Thulamela Municipality, Vhembe District, Limpopo Province, South Africa. Vhembe District covers a geographical area that is predominantly rural. Information about socio-demographic characteristics, household income, disease family history, and level of education of parents was obtained using a researcher-administered questionnaire.

### Anthropometric measurements

Body height was measured, without wearing shoes, by trained fieldworkers with an accuracy of 0.1 cm, using a portable Harpenden stadiometer (Holtain Ltd., Crymych, UK). Bodyweight was measured with participants wearing light clothing using a calibrated electronic scale (SECA, Birmingham, UK) with an accuracy of 0.1 kg. Body mass index (BMI) in kg/m^2^ was determined and classified according to International Obesity Task Force (IOTF) cut-off points for children and adolescents [22]. BMI was used as a proxy for overall obesity.

Waist circumference was measured using an inelastic flexible measuring tape. The participants were wearing light clothing. The researchers located the top of the right iliac crest, the highest point of the hip bone on the right side. The measuring tape was in a horizontal plane around the abdomen at the level of the iliac crest. The measurements were recorded to the nearest 0.1 cm and waist circumference for adolescents 18 years and below was classified according to age and sex [23]. For adolescents above 18 years, the WHO cut-off points of WC were used [24].

The participant stood erect with arms at the side and feet together. The measurement was taken at the point yielding the maximum circumference over the buttocks, with the tape held in a horizontal plane, touching the skin but indenting the soft tissue [25]. Waist and hip circumferences were measured to determine Waist-to-hip-ratio (WHR). WHR was calculated and the WHO cut-off points were used [24]. Waist circumference was used as a proxy to determine android obesity.

### Biochemical measurements

A professional nurse was responsible for collecting blood samples. One participant was brought to the room at a time. The participant’s skin was cleaned with alcohol-soaked gauze pads at the tip of the finger. The participant’s finger was pricked, and the blood drops were allowed fall on the test panel of a Cardio-check device. A Cardio-check device (model CE 0197) was used to determine total cholesterol (TC), high density lipoprotein cholesterol (HDL-C), low density lipoprotein cholesterol (LDL-C), triglycerides (TG), and glucose level. TC, LDL-C, HDL.C and TG were classified according to age and gender lipoprotein for adolescents [26]. Glucose level was classified according to laboratory standards.

### Clinical assessment: Blood pressure (BP)

A professional nurse was responsible for taking blood pressure readings using standard procedures. BP reading was taken using a digital automated device OMRO (model CE0197). Each participant sat quietly for 5-10 minutes after their arm was placed at heart level and BP was measured at least 3 times in 5 minutes intervals. If BP varied in these determinations by greater than 10mmHg, 3 additional trials were performed to measure systolic and diastolic blood pressure. BP for adolescents less than 18 years were classified according to sex, age, and height [27]. For adolescents ≥18 years BP was classified according to American Heart Association (AHA) for adults [28].

### Assessment of dietary Intake

A quantified food frequency questionnaire (FFQ) was used to assess the dietary intake of the study participants. The researchers gave the adolescents a pile of pictures with food items and instructed them to put pictures of food items he/she ate in the past week on the right hand and the ones they did not eat on the left-hand side. From the selected food items, adolescents were then asked about how many times per week and per day they consumed the foods. The standard portion sizes for food items were included in the FFQ. With both the frequency of consumption and the standard portion sizes the total portion sizes were calculated. The food models and the line drawing from the DAEK manual assisted the adolescents to estimate the portion sizes. The data gathered were analysed using the Food Finder computer software (version 1.1.3) to assess the dietary intake. The aim of the analysis was to determine the amount of each nutrient and energy consumed per day. These nutrients included carbohydrates, lipids, proteins, minerals, and vitamins.

### Physical activity

A physical activity questionnaire by Sharkey and Gaskill [29] was adopted to assess the physical activity level. A questionnaire included physical activity patterns in and around the house, travelling to school and recreations that participants engaged in. Information such as sedentary lifestyle, after-school activities, time spent on television viewing, and sleep duration frequency was included in the questionnaire. Table 1 indicate the interpretation of physical activity indices. The following formula was used to calculate physical activity indices:

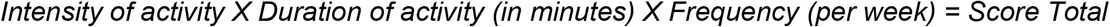

**Table 1:** Interpretation of the physical activity indices scores [29].

| Evaluation of activity score |  |  |
| --- | --- | --- |
| Score | Evaluation | Activity category |
| 81 to 100 | Very active lifestyle | High |
| 60 to 80 | Active and healthy | Very good |
| 40 to 59 | Acceptable but could be better | Fair |
| 20 to 39 | Not good enough | Poor |
| Under 20 | Sedentary |  |

Adopted from [29].

### Statistical analysis

Data were checked for completeness and consistency. Data were entered, cleaned, and analysed using SPSS (IBM Corporation, USA) version 26 statistical package software. Descriptive statistics like frequencies and proportions were used to summarize the data. Shapiro Wilk and Kolmogorov Smirnov test were used to test for normality. The data was homogenous. ANOVA was used to determine the differences between BMI classifications. Logistic regression was used for bivariate and multivariate analyses to calculate unadjusted and adjusted odds ratios (AOR) for determinants of obesity. Statistical significance was set at p < 0.05.

### Ethics approval

The study was approved by the Research and Ethics Committee of the University of Venda and the Ethical clearance certificate was issued (SHS/17/NUT/03/1506). Permission to conduct the study was granted by the Provincial and District Departments of Basic Education. No participant could participate in the study without signed informed consent by parents and an assent form by participants after a full and adequate explanation of the study. The data obtained from the study were kept on a computer database in such a manner that it maintains the participants’ confidentiality (a code was assigned to each participant for these purposes).

## RESULTS

### Demographic characteristics

The characteristics of the study participants are indicated in Table 2. The mean age of the study participants was 16.56±2.10. Majority were of Vha-Venda ethnic group. The frequency of buying food at school was more prevalent in the overweight and obese group for both boys and girls. Majority of the guardians had secondary education. Parents with secondary school education had children with the highest prevalence of overweight and obesity. Learners whose parents/guardians receive a salary or wages had a higher prevalence of overweight and obesity.

**Table 2.** Characteristics of the study participants.

| Variables | Total | Boys |  |  |  | Girls |  |  |  |
| --- | --- | --- | --- | --- | --- | --- | --- | --- | --- |
|  |  | Normal | Overweight | Obese | P-value | Normal | Overweight | Obese | P-value |
| Adolescent (mean ±SD) |  |  |  |  |  |  |  |  |  |
| Age in years | 16.56±2.10 | 16.89±2.08 | 17.14±2.27 | 17.00±1.66 | 0.945 | 16.25±2.16 | 16.68±1.79 | 16.32±2.03 | 0.639 |
| Grade n(%) |  |  |  |  |  |  |  |  |  |
| Grade 8 | 68 (18.1%) | 25 (18.8%) | 1 (14.3%) | 1 (11.1%) |  | 33 (20.4%) | 1 (5.3%) | 7 (15.2%) |  |
| Grade 9 | 79 (21.0%) | 28 (21.1%) | 2 (28.6%) | 3 (33.3%) |  | 27 (16.7%) | 6 (31.6%) | 13 (28.3%) |  |
| Grade 10 | 88 (23.4%) | 36 (27.1%) | 2 (28.6%) | 3 (33.3%) |  | 36 (22.2%) | 5 (26.3%) | 6 (13.0%) |  |
| Grade 11 | 73 (19.4%) | 21 (15.8%) | 1 (14.3%) | 1 (11.1%) |  | 35 (21.6%) | 5 (26.3%) | 10 (21.7%) |  |
| Grade 12 | 68 (18.1%) | 23 (17.3%) | 1 (14.3%) | 1 (11.1%) |  | 31 (19.1%) | 2 (10.5%) | 10 (21.7%) |  |
| Ethnics group n(%) |  |  |  |  |  |  |  |  |  |
| Tsonga | 5 (1.7%) | 2 (1.5%) | 0 | 0 |  | 3 (1.9%) | 0 | 0 |  |
| Venda | 364 (96.8%) | 130 (97.7) | 7 (100%) | 9 (100%) |  | 155 (95.7%) | 18 (94.7%) | 45 (97.8%) |  |
| Pedi | 7 (1.9%) | 1 (0.8%) | 0 | 0 |  | 4 (2.5%) | 1 (5.3%) | 1 (2.2%) |  |
| Pocket money n(%) |  |  |  |  |  |  |  |  |  |
| None | 10 (2.7%) | 4 (3.0%) | 1 (14.3%) | 0 |  | 5 (3.1%) | 0 | 0 |  |
| <R5 | 34 (9.0%) | 4 (3.0%) | 0 | 1 (11.1%) |  | 20 (12.3%) | 2 (10.5%) | 7 (15.2%) |  |
| R5-R10 | 252 (67.0%) | 89 (66.9%) | 4 (57.1%) | 7 (77.8%) |  | 112 (69.1%) | 9 (47.4%) | 31 (67.4%) |  |
| R10-R15 | 63 (16.8%) | 25 (18.8%) | 2 (28.6%) | 1 (11.1%) |  | 22 (13.6%) | 8 (42.1%) | 5 (10.9%) |  |
| R15-R20 | 16 (4.3%) | 10 (7.5%) | 0 | 0 |  | 3 (1.9%) | 0 | 3 (6.5%) |  |
| >R20 | 1 (0.3%) | 1 (0.8%) | 0 | 0 |  | 0 | 0 | 0 |  |
| Lunch box n(%) |  |  |  |  |  |  |  |  |  |
| 1-2 times | 24 (6.4%) | 6 (4.5%) | 0 | 1 (11.1%) |  | 14 (8.6%) | 2 (10.5%) | 1 (2.2%) |  |
| 3-4 times | 37 (9.8%) | 4 (3.0%) | 0 | 0 |  | 24 (14.8%) | 2 (10.5%) | 7 (15.2%) |  |
| Everyday | 30 (8.0%) | 7 (5.3%) | 0 | 0 |  | 15 (9.3%) | 4 (21.1%) | 4 (8.7%) |  |
| Never | 285 (75.8%) | 116 (87.2%) | 7 (100%) | 8 (88.9%) |  | 109 (67.3%) | 11 (57.9%) | 34 (73.9%) |  |
| Buying food at school n(%) |  |  |  |  |  |  |  |  |  |
| 1-2 times | 49 (13.0%) | 21 (15.8%) | 0 | 2 (22.2%) |  | 19 (11.7%) | 3 (15.8%) | 4 (8.7%) |  |
| 3-4 times | 105 (27.9%) | 36 (27.1%) | 2 (28.6%) | 4 (44.4%) |  | 44 (27.2%) | 4 (21.1%) | 15 (32.6%) |  |
| Everyday | 195 (51.9%) | 68 (51.1%) | 4 (57.1%) | 2 (22.2%) |  | 85 (52.5%) | 10 (52.6%) | 26 (56.5%) |  |
| Never | 27 (7.2%) | 8 (6.0%) | 1 (14.3%) | 1 (11.1%) |  | 14 (8.6%) | 2 (10.5 %) | 1 (2.2%) |  |
| Source of income of parents |  |  |  |  |  |  |  |  |  |
| Salary | 102 (27.1%) | 36 (27.1%) | 3 (42.9%) | 4 (44.4%) |  | 41 (25.3%) | 3 (15.8%) | 15 (32.6%) |  |
| Pension grant | 68 (18.1%) | 31 (23.3%) | 1 (14.3%) | 1 (11.1%) |  | 24 (14.8%) | 5 (26.3%) | 6 (13.0%) |  |
| Child support grant | 64 (17.0 %) | 21 (15.8%) | 0 | 1 (11.1%) |  | 31 (19.1%) | 5 (26.3%) | 6 (13.0%) |  |
| Wages | 137 (36.4%) | 44 (15.8%) | 3 (42.9%) | 3 (33.3%) |  | 63 (38.9%) | 6 (31.6%) | 18 (39.1%) |  |
| Disability grant | 4 (1.1%) | 0 | 0 | 0 |  | 3 (1.9%) | 0 | 1 (2.2%) |  |
| Pension and child support grant | 1 (0.3%) | 1 (0.8%) | 0 | 0 |  | 0 | 0 | 0 |  |
| Marital status of parents n(%) |  |  |  |  |  |  |  |  |  |
| Married | 110 (29.3%) | 38 (28.6%) | 2 (28.6%) | 4 (44.4%) |  | 44 (27.2%) | 5 (26.3%) | 17 (37.0%) |  |
| Divorced | 39 (10.4%) | 14 (10.5%) | 1 (14.3%) | 0 |  | 17 (10.5%) | 1 (5.3%) | 6 (13.0%) |  |
| Windowed | 41 (10.9%) | 16 (12.0%) | 1 (14.3%) | 1 (11.1%) |  | 16 (9.9%) | 16 (9.9%) | 4 (8.7%) |  |
| Living together | 100 (26.6%) | 38 (28.6%) | 1 (14.3%) | 2 (22.2%) |  | 44 (27.2%) | 44 (27.2%) | 13 928.3%) |  |
| Single | 86 (22.9%) | 27 (20.3%) | 2 (28.6%) | 2 (22.2%) |  | 41 (25.3%) | 41 (25.3%) | 6 (13.0%) |  |
| **Highest level of education of parents n(%)** |  |  |  |  |  |  |  |  |  |
| Never attended school | 10 (2.7%) | 6 (4.5%) | 0 | 0 |  | 4 (2.5%) | 0 | 0 |  |
| Primary school | 46 (12.2%) | 19 (14.3%) | 2 (28.6%) | 2 (22.2%) |  | 14 (8.6%) | 4 (21.1%) | 5 (47.4%) |  |
| Secondary school | 264 (70.2%) | 87 (65.4%) | 3 (42.9%) | 7 (77.8%) |  | 121 (74.7%) | 9 (47.4%) | 37 (80.4%) |  |
| Tertiary | 56 (14.9%) | 21 (15.8%) | 2 (28.6%) | 0 |  | 23 (14.2%) | 6 (31.6%) | 4 (8.7%) |  |
| **Type of employment of parents n(%)** |  |  |  |  |  |  |  |  |  |
| Government | 53 (14.1%) | 21 (15.8%) | 1 (14.3%) | 0 |  | 21 (13.0%) | 3 (15.8%) | 7 (15.2%) |  |
| Private company | 136 (36.2%) | 43 (32.3%) | 4 957.1%) | 6 (66.7%) |  | 63 (38.9%) | 3 (15.8%) | 17 (37.0) |  |
| Self employed | 59 (15.7%) | 23 (17.3%) | 0 | 1 (11.1%) |  | 24 (14.8%) | 4 (21.1%) | 7 (15.2%0 |  |
| Unemployed | 128 (34.0%) | 46 (34.6%) | 2 (28.6%) | 2 (22.2%) |  | 54 (33.3%) | 9 (47.4%) | 15 (32.6%) |  |
| **Number of people in household (HH) n(%)** |  |  |  |  |  |  |  |  |  |
| 1-2 | 31 (8.2%) | 14 (10.5%) | 1 (14.3%) | 2 (22.2%) |  | 9 (5.6%) | 2 (10.5%) | 3 (6.5%) |  |
| 3-4 | 122 (32.4%) | 45 (33.8%) | 2 (28.6%) | 2 (22.2%) |  | 54 (33.3%) | 4 (21.1%) | 15 (32.6%) |  |
| 5-6 | 146 (38.8%) | 52 (39.1%) | 2 (28.6%) | 3 (33.3%) |  | 60 (37%) | 8 (42.1%) | 21 (45.7%) |  |
| 7-8 | 63 (16.8%) | 19 (14.3%) | 2 (28.6%) | 2 (22.2%) |  | 29 (17.9%) | 4 (21.1%) | 7 (15.2%) |  |
| 9 and above | 14 (3.7 | 3 (2.3%) | 0 | 0 |  | 10 (6.2%) | 1 (5.3%) | 0 |  |

### Anthropometry, clinical assessment, and biochemical measurements of study participants

The prevalence of abdominal obesity among boys and girls was 22.2% and 32.6% respectively. The high prevalence of substantially increased WHR was seen in overweight boys and obese girls. Among the classified as obese, only 13% had hypertensive SBP (Table 3).

**Table 3.** Anthropometry, clinical assessment, and biochemical measurements of study participants.

| Variables | Total | Boys |  |  | Girls |  |  |
| --- | --- | --- | --- | --- | --- | --- | --- |
|  |  | Normal | Overweight | Obese | Normal | Overweight | Obese |
| WC n (%) |  |  |  |  |  |  |  |
| Normal | 354<br>(94.1%) | 133<br>(100%) | 7 (100%) | 7 (77.8%) | 157<br>(96.9%) | 19 (100) | 31 (67.4%) |
| Abdominal obesity | 22<br>(5.9%) | 0 | 0 | 2<br>(22.2%) | 5 (3.1%) | 0 | 15 (32.6%) |
| WHR n (%) |  |  |  |  |  |  |  |
| Normal | 344<br>(91.5%) | 122<br>(91.7%) | 6 (85.7%) | 8<br>(89.9%) | 156<br>(96.3%) | 19 (100%) | 33 (71.7%) |
| Substantially increased | 32<br>(8.5%) | 11 (8.3%) | 1 (14.3%) | 1<br>(11.1%) | 6 (3.7%) | 0 | 13 (28.3%) |
| SBP n (%) |  |  |  |  |  |  |  |
| Normal | 301<br>(80.1%) | 90<br>(67.7%) | 6 (85.7%) | 8<br>(88.9%) | 149<br>(92.0%) | 16 (84.2%) | 36 (78.3%) |
| Pre-hypertensive | 54<br>(14.4%) | 35<br>(26.3%) | 1 (14.3%) | 1<br>(11.1%) | 8 (4.9%) | 2 (10.5%) | 4 (8.7%) |
| Hypertensive | 20<br>(5.3%) | 8 (6.1%) | 0 | 0 | 5 (3.1%) | 1 (5.3%) | 6 (13.0%) |
| DBP n (%) |  |  |  |  |  |  |  |
| Normal | 310<br>(82.4%) | 118<br>(88.7%) | 5 (71.4%) | 7<br>(77.8%) | 135<br>(83.3%) | 15 (78.9%) | 30 (65.2%) |
| Pre-hypertensive | 37<br>(9.8%) | 13 (9.8%) | 2 (28.6%) | 1<br>(11.1%) | 14 (8.6%) | 0 | 7 (15.2%) |
| Hypertensive | 29<br>(7.7%) | 2 (1.5%) | 0 | 1<br>(11.1%) | 13 (8.0%) | 4 (21.1%) | 9 (19.6%) |
| BGL n (%) |  |  |  |  |  |  |  |
| Normal | 366<br>(97.3%) | 131<br>(98.5%) | 7 (100%) | 9 (100%) | 155<br>(95.7%) | 19 (100%) | 45 (97.8%) |
| Pre-Diabetic | 8 (2.1%) | 2 (1.5%) | 0 | 0 | 6 (3.7%) | 0 | 0 |
| Diabetic | 2 (0.5%) | 0 | 0 | 0 | 1 (0.6%) | 0 | 1 (2.2%) |
| TC n (%) |  |  |  |  |  |  |  |
| Normal | 367<br>(97.6%) | 133<br>(100%) | 6 (85.7%) | 9 (100%) | 157<br>(96.9%) | 18 (94.7%) | 44 (95.7%) |
| Borderline high | 8 (2.1%) | 0 | 1 (14.3%) | 0 | 4 (2.5%) | 1 (5.3%) | 2 (4.3%) |
| High | 1 (0.3%) | 0 | 0 | 0 | 1 (0.6%) | 0 | 0 |
| LDL n (%) |  |  |  |  |  |  |  |
| Normal | 370<br>(98.4%) | 133<br>(100%) | 7 (100%) | 9 (100%) | 158<br>(97.5%) | 19 (100%) | 44 (95.7%) |
| Above normal | 5 (1.3%) | 0 | 0 | 0 | 3 (1.9%) | 0 | 2 (4.3%) |
| Borderline high | 1 (0.3) | 0 | 0 | 0 | 1 (0.6%) | 0 | 0 |
| HDL n(%) |  |  |  |  |  |  |  |
| Low | 160<br>(42.6%) | 69<br>(51.9%) | 6 (85.7%) | 3<br>(33.3%) | 52<br>(32.2%) | 6 (31.6%) | 24 (52.2%) |
| Normal | 173<br>(46.0%) | 56<br>(42.1%) | 1 (33.3%) | 6<br>(66.7%) | 83<br>(51.2%) | 7 (36.8%) | 20 (43.5%) |
| Protective | 43<br>(11.4%) | 8 (6.0%) | 0 | 0 | 27<br>(16.7%) | 6 (31.6%) | 2 (4.3%) |
| TRIG n(%) |  |  |  |  |  |  |  |
| Normal | 341<br>(90.7%) | 125<br>(94%) | 7 (100%) | 6<br>(66.7%) | 150<br>(92.6%) | 16 (84.2%) | 37 (80.4%) |
| Borderline high | 17<br>(4.5%) | 6 (4.5%) | 0 | 2<br>(22.2%) | 6 (3.7%) | 0 | 3 (6.5%) |
| High | 18<br>(4.8%) | 2 (1.5%) | 0 | 1<br>(11.1%) | 6 (3.7%) | 3 (15.8%) | 6 (13.0%) |

### BMI classification of the study participants

The mean BMI of boys was 20.81±3.45 and that of girls was 23.3±4.90. There was a significant difference between boys and girls in terms of BMI P-value was less than 0.005. The results of the study population show that the prevalence of overall combined overweight/obesity is 21.5%. The prevalence of overweight and obesity was higher in girls than in boys (Figure 1).

**Figure 1:**
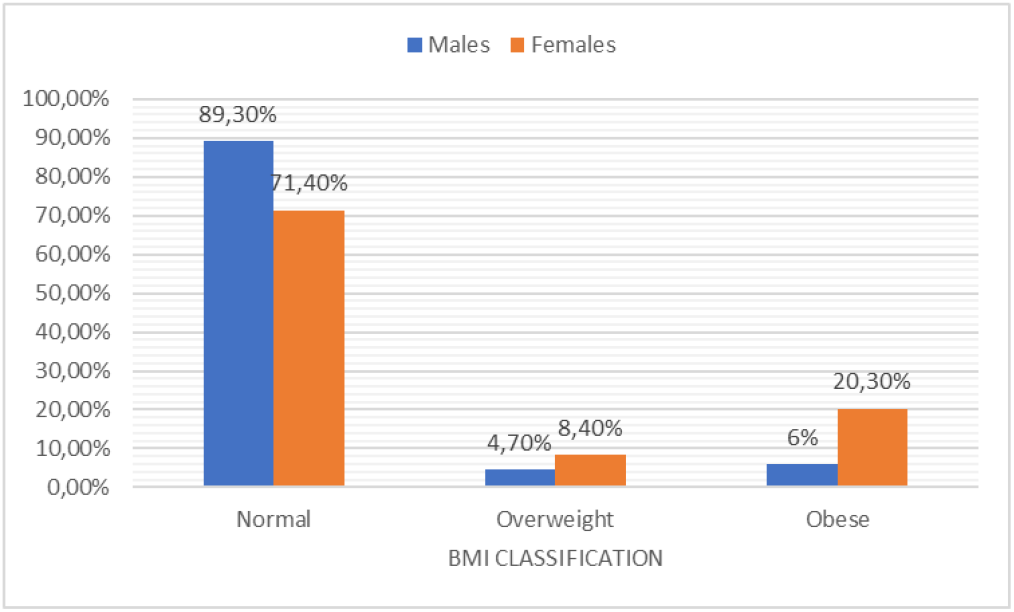
BMI classification

### Determinants of obesity

The determinants of obesity are shown in Table 4. Gender (female) (β=0.32, p=0.018, 95%CI); age (β=1.28, p=0.015, 95%CI); source of income for parents who earn salary or wages (β=3.25, p=0.008, 95%CI) and systolic blood pressure (β=1.04, p=0.01, 95%CI) were associated with obesity.

**Table 4.** Determinants of obesity.

| Determinates | Unadjusted Odds ratio (OR) | Adjusted OR | 95%-CI | p-value |
| --- | --- | --- | --- | --- |
| Gender | -1.145 | 0.32 | (0.12-0.82) | 0.018** |
| Age | 0.246 | 1.28 | (1.05-1.56) | 0.015** |
| Source of Income (salary or wages) | 1.180 | 3.25 | (1.36-7.78) | 0.008*** |
| Moderate exercise | -0.946 | 0.39 | (0.12-1.2) | 0.107* |
| Systolic BP | 0.041 | 1.04 | (1.02-1.07) | 0.01** |
| LDL | 1.387 | 0.25 | (0.09-0.67) | 0.006*** |
| HDL | -2.454 | 0.09 | (0.02-0.34) | 0.000*** |
| TC/HDL | 1.383 | 3.99 | (2.17-7.31) | 0.000*** |
| Energy (KJ) | 0.006 | 0.99 | (0.99-1.00) | 0.047** |
| Total fat (g) | 0.217 | 1.24 | (0.99-1.57) | 0.067* |
| Total trans FA(g) | 0.635 | 1.89 | (1.04-3.43) | 0.037** |
| Carbohydrate, avai.(g) | 0.111 | 1.12 | (1.00-1.25) | 0.049** |
| Added sugar | 0.022 | 0.98 | (0.96-1.00) | 0.063* |

\*\*\* Highly significant (at alpha = 0.01), \*\* Significant (at alpha = 0.05), \* Barely significant (at alpha=0.10)

### Comparison of means of variables

Table 5 below compares the means of variables. For WC there was a significant difference between BMI categories in both males and females. Among females, there was a significant difference between BMI categories in W/HR, SBP, HDL and TCHDL.

**Table 5.** Comparisons of means of variables.

| Variables | Total (mean ± SD) | Boys (mean ± SD) | | | | Girls (mean ± SD) | | | |
| --- | --- | --- | --- | --- | --- | --- | --- | --- | --- |
|  |  | Normal | Overweight | Obese | p-value | Normal | Overweight | Obese | p-value |
| WC (cm) | 71.73±10.23 | 67.31±5.90 | 77.49±7.72 | 89.22±9.44 | 0.000 | 68.95±6.00 | 76.95±5.56 | 88.22±13.02 | 0.000 |
| W/HR | 0.78±0.77 | 0.79±0.94 | 0.82±0.10 | 0.81±0.05 | 0.472 | 0.77±0.05 | 0.77±0.04 | 0.81±0.08 | 0.000 |
| SBP (mmHg) | 110.78±13.41 | 116.02±12.92 | 117.29±9.49 | 125.06±12.90 | 0.125 | 105.24±11.26 | 109.02±14.73 | 112.16±13.40 | 0.002 |
| DBP (mmHg) | 71.42±9.14 | 69.59±8.50 | 74.57±3.33 | 71.5±8.19 | 0.257 | 71.75±9.28 | 70.63±12.27 | 75.80±8.91 | 0.027 |
| BGL (mmol/L) | 4.24±0.84 | 4.15±0.66 | 4.33±0.82 | 4.57±0.67 | 0.161 | 4.26±0.78 | 4.12±0.59 | 4.43±1.43 | 0.388 |
| LDL | 0.62±0.82 | 0.36±0.65 | 0.64±0.82 | 1.09±1.05 | 0.007 | 0.73±0.86 | 0.64±0.756 | 0.91±0.91 | 0.390 |
| HDL | 1.14±0.33 | 1.07±0.29 | 0.93±0.16 | 1.12±0.26 | 0.403 | 1.22±0.35 | 1.30±0.33 | 1.04±0.29 | 0.004 |
| TCHDL | 1.13±1.41 | 0.68±1.19 | 0.90±1.55 | 1.86±1.81 | 0.025 | 1.23±1.36 | 1.09±1.29 | 2.38±1.67 | 0.003 |

Waist Circumference (WC), Waist to Hip Ratio (WHR), Systolic Blood Pressure (SBP), Diastolic Blood Pressure (DBP), Blood Glucose Level (BGL), Low-Density Lipoprotein (LDL), High-Density Lipoprotein (HDL) Total Cholesterol High-Density Lipoprotein Ratio (TCHDL)

### Comparison of means of nutrient intake

The comparison of means of nutrient intake is discussed in Table 6. The result of the study shows that boys who were in obese and normal category did not meet daily energy requirements, whereas obese girls were consuming more energy. Boys in the obese category were consuming low Total fats as compared to boys in the normal and overweight classification and girls in all classifications who were consuming more than recommended daily total fats intake. When it comes to Total fibre only obese boys who do not meet daily fibre intake. All adolescents were consuming more than the daily recommendations in carbohydrates.

**Table 6.** Comparisons of means of nutrients intake.

| Variables | Total (mean ±SD) | Boys (mean ±SD) |  |  |  |  | Girls (mean ±SD) |  |  |  |  |
| --- | --- | --- | --- | --- | --- | --- | --- | --- | --- | --- | --- |
|  |  | RDIs/<br>AI/AM<br>DRs | Normal | Overweight | Obese | p-value | RDIs/<br>AI/AM<br>DRs | Normal | Overweight | obese | p-value |
| Energy (kj) | 10470.05±6480.32 | 12552<sup>a</sup> | 11114.66±5514.04 | 12594.25±4498.49 | 9100.02±3287.13 | 0.413 | 9205<sup>a</sup> | 9968.92±7325.81 | 9290.67±3943.81 | 10676.59±7341.29 | 0.743 |
| Total protein (g) | 62.63±36.29 | 52<sup>a</sup> | 66.31±32.71 | 74.69±2567 | 55.19±17.43 | 0.457 | 46<sup>a</sup> | 60.01±40.03 | 57.99±24.74 | 62.01±40.02 | 0.921 |
| Total fat (g) | 49.79±28.28 | 20-35<sup>c</sup> | 50.67±26.21 | 55.74±18.72 | 34.76±9.27 | 0.160 | 20-35<sup>c</sup> | 48.74±26.27 | 42.89±16.42 | 55.43±43.48 | 0.245 |
| Total trans FA (g) | 1.39±1.32 | - | 1.31±1.41 | 1.54±1.65 | 0.98±0.67 | 0.700 | - | 1.44±1.27 | 1.20±0.96 | 1.52±1.44 | 0.655 |
| Cholesterol (mg) | 176.28±181.54 | - | 180.59±179.35 | 190.56±127.46 | 124.29±94.23 | 0.631 | - | 167.79±175.97 | 174.93±150.79 | 199.95±233.89 | 0.590 |
| Total dietary fibre (g) | 32.19±22.32 | 38<sup>b</sup> | 35.09±18.56 | 43.83±20.78 | 30.90±12.48 | 0.363 | 26<sup>b</sup> | 30.10±26.24 | 27.66±13.74 | 31.53±21.13 | 0.844 |
| Carbohydrate, avail (g) | 412.39±278.73 | 130<sup>a</sup> | 442.49±230.59 | 501.49±205.22 | 373.98±151.95 | 0.524 | 130<sup>a</sup> | 390.53±325.74 | 368.06±172.79 | 414.65±288.76 | 0.836 |
| Added sugar (g) | 27.09±27.23 | - | 24.99±26.48 | 39.27±35.59 | 10.89±7.22 | 0.101 | - | 28.62±24.72 | 29.66±35.54 | 28.06±33.91 | 0.978 |

a- Recommended Dietary Allowance (RDA), b- Adequate Intake (AI), c- Acceptable Macronutrients Distribution Range (AMDR)

### Physical activity

Majority of overweight and obese boys and girls led a sedentary lifestyle. The results show that the prevalence of a sedentary lifestyle was high in overweight and obese girls than in boys. At least a few percentages of obese boys had a very active lifestyle as compared to girls (Figure 2).

**Figure 2.**
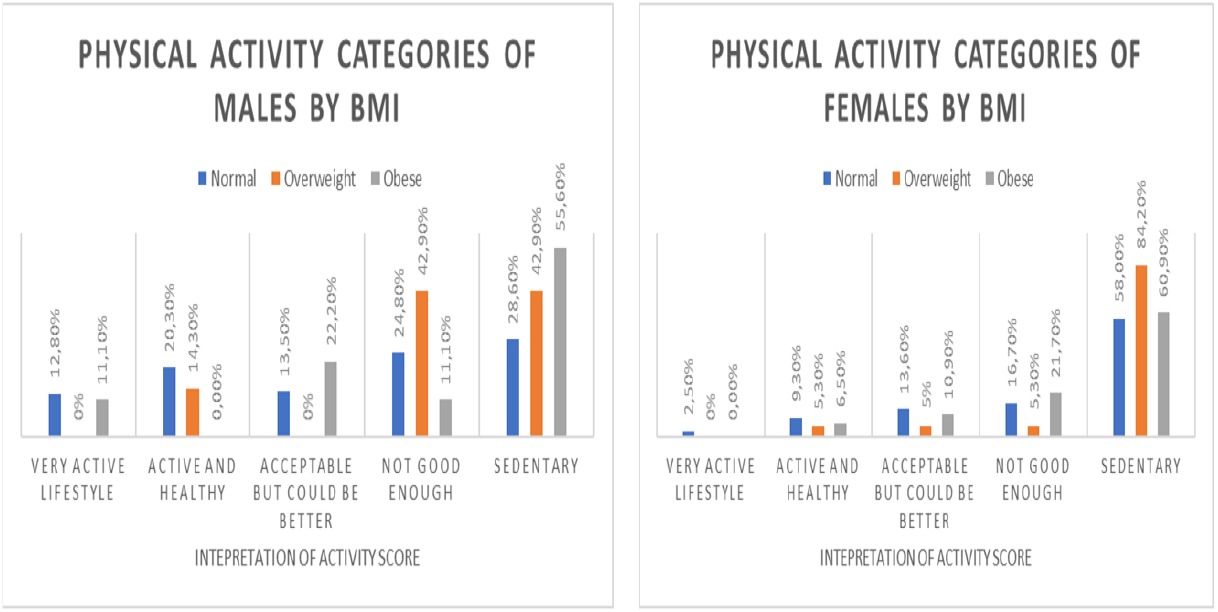
Physical activity categories of males and females by BMI. Physical activity

## DISCUSSION

The aim of the study was to describe the determinants of obesity among rural secondary school learners in Thulamela Municipality, Vhembe District, Limpopo province, South Africa. The findings showed the prevalence of overweight and obesity to be 21.5%. The prevalence of overweight and obesity was lower than that of 35% by Debeila et al. (2021) [30] in rural high schools in Limpopo province South Africa. It was also lower than the prevalence found in the provincial dietary intake study conducted in Gauteng (overweight 27.7% vs obese 39.1%) and the Western Cape (overweight 20.4% vs obese 50.6%) [31]. The overall prevalence of overweight and obesity in this study is lower than the national range of 8.6% to 27.0% among adolescents aged 15 to 19 years reported by UNICEF 2018 [32]. The prevalence of overall overweight and obesity is lower than that of other studies conducted in urban and rural settings in South Africa. Recently, a prevalence of 22.9% of overweight/obesity among adolescents in urban high schools in the Western Cape in South Africa has been reported [33]. The prevalence of overweight and obesity reported in this study is lower compared to studies conducted by Negash et al. (2017) and UICEF (2018) [33,32] in South Africa. Although the prevalence in the present study is lower than that of other studies, it is evident that overweight and obesity is increasing in the rural population [34].

The result of the current study showed that the prevalence of overweight and obesity was higher in females (28.7%) than in males (10.7%). The results are comparable with several studies done in South Africa [33,18,20]. However, the combined prevalence of overweight and obesity reported in girls is almost the same as that of Negash et al. (2017) [33] with the prevalence of overweight and obesity in girls being 28.8%. Physiologically girls require less energy than boys [35]. On a behavioural level, girls are reported to be more attentive to food and its effect on health and weight control than boys [36]. In addition, adolescent girls are less likely to meet physical activity recommendations than boys [37].

In addition to the overall overweight and obesity prevalence, the results found a prevalence of abdominal obesity indicated by elevated WC (5.9%) and elevated WHR (8.5%). As seen in the results, abdominal obesity was high in girls than in boys. The result suggests that girls are significantly affected by abdominal obesity. Sekokotla et al. (2017) [38] reported abdominal obesity among adolescents in South Africa. Studies done amongst students in Ghana by [39,40] Mogre et al. (2014) and Mogre et al. (2015) highlighted higher odds of abdominal obesity amongst females compared to males. Adolescent females are twice as likely to be abdominal obese than males based on WC and WHR indicators. The prevalence reported in this study highlight a public health implication. Abdominal obesity in children and adolescents is strongly associated with metabolic risk factors compared to overall obesity [41,42].

The results of the study show that majority of the participants were leading a sedentary lifestyle. Females were less active compared to males. This is not surprising since studies have reported that girls are less likely to be physically active as compared to boys [37,43-47]. The WHO report ranks physical inactivity as the fourth leading cause of global mortality, with responsibility for 6% of deaths worldwide [48,49]. Kallio et al. (2021) [50] reported that a physical inactivity lifestyle is associated with cardiometabolic risk factors. Additionally, Tremblay et al. (2011) [51] found that higher sedentary behaviour is associated with adverse health outcomes.

The results of the study suggest that being female increased the odds of being obese. Studies [52,33] tracking the determinants of obesity arrived at similar conclusions where it was reported being female was a determinant of obesity and females were twice likely to be obese than males. Female adolescents tend to have higher BMI as a result of rapid growth and early sexual maturity [53].

An increase in age was associated with the odds of being obese. Similarly, the study conducted in India reported that adolescents above 14 years and older had 2.09 times more odds of being obese [54]. Studies tracking the prevalence of obesity in SA indicate that the prevalence of obesity increases with the age, peaking at age 12 years and declining thereafter [55,17]. This may be due to the increase in adipose tissue and overall body weight in adolescents during puberty [56]. According to Kimani-Murage et al. (2010) [4], the risk of obesity increases with sexual maturation, indicating a higher risk as the adolescent transitions to adulthood.

In the current study parent who earns salary or wages as a source of income was strongly associated with obesity. The results of the current study show that parent source of income is a significant determinant of obesity among adolescents. The results of the study are consistent with the findings of Choukem et al. [57] where high SES was strongly associated with obesity among children, with almost 2.5 times more likely to be obese than those with low SES. Concurrently a study conducted on Moroccan adolescents, reported that high family income was a determinant of obesity [58]. Money gives children a certain degree of autonomy in purchasing and consumption [59,60] some of which entail health risks [61] such as smoking and substance abuse [62-66]. Studies from US, Europe, India, Korea and Vietnam suggest that pocket money is a potential risk factor for child’s unhealthy eating and thus overweight and obesity [59,60,67-70].

The odds of obesity increased in participants who do not engage in moderate exercise. As observed in the current study, many of the females were not physically active. Therefore, it can be deduced that females are at greater risk of being obese. Ahmad et al. [53] maintain that girls engage in less physical activity and sport, as compared to boys of great concern, is the cultural influence that girls do not move around than boys leading to physical inactivity and eventually the development of obesity [3]. Studies, [71,46] reported a decline in physical activity among adolescents. Furthermore, physical inactivity and increased sedentary time play a role in the development of obesity [71].

The results of the study revealed that 1 mmHg of SBP was associated with an increase in the odds of being obese. Concurrently, Ghomari-Boukhatem et al. and Negash et al. [72,33] reported that hypertension increases the odds of being obese among adolescents. Confirming the findings of the current study, are studies conducted in SA where SBP was associated with obesity [73,74]. The association of hypertension and obesity among adolescents may highlight those cardio-metabolic comorbidities which do not only occur in adults but may develop in children and adolescents, particularly in the presence of obesity [33]. The relationship between hypertension and obesity decrees effects which may be attributable to the nutrition transition which results in sedentary living and overfeeding [73].

The present study reveals that high TC, LDL, and low HDL as determinants of obesity. This is similar to the study conducted by Ghomari-Boukhatem et al. [73] which reported elevated TC, LDL-C, TG and low HDL-C as risk factors for obesity. Furthermore, a study conducted in the USA among children and adolescents shows that youth with obesity had a higher prevalence of high TC and low HDL-C than those with normal weight [77]. Bibilon et al. [76] stated that obese children were associated with the existence of at least one abnormal lipoprotein concentration. Studies, [77-80] reported that abnormalities in these lipoproteins as the strongest risk factor for CVD. Miller et al. and D’Adamo et al. [77,81] maintain that atherogenesis is associated with lipoprotein and obesity.

High consumption of energy, total fats, total trans fats, carbohydrates and added sugar were reported as determinants of obesity in the current study. Similarly, Appannah et al. [82] reported that energy-dense and high fat intake is associated with cardiometabolic risk factors insulin resistance and obesity among adolescents. Allioua et al. [83] also reported that high consumption of fats was associated with obesity among adolescents. A high energy intake is a major risk factor for obesity in children and adults [17]. In addition, high energy intake together with high total fats, high saturated fats, high carbohydrates, high added sugar and low fibre intake has been classified as western diet which contributes to the development of chronic diseases [17]. Studies [19,84,85] show that a high intake of fats and sugar increases the risk of childhood obesity, as well as non-communicable diseases such as type 2 diabetes and CVD, later in life. Low fibre intake was observed in obese boys, while other adolescents were meeting the daily recommendation. Dreher. [86] reported that fibre is one of the under-consumed nutrients and low intake increases weight gain over time. Studies, [86,87] show that high fibre intake to be associated with lower anthropometric indices including weight and WC.

Adolescents are nutritionally vulnerable, they frequently make poor food choices [85]. Furthermore, school children spend more time at school and most vendors sell energy-dense food such as crisps, fat cakes, French fries, sweets, and carbonated sweet drinks [88]. The association between high total fats, trans fats and carbohydrate intake and obesity in the present study suggests that there is a peak in the nutritional transition and weight status in rural communities.

## LIMITATIONS

The study has several limitations. The study was cross-sectional in nature using adolescents from the different geographical areas within Thulamela municipality. The number of adolescents included in the study was small and generalisability within this group would be limited. The study did not determine the BMI and dietary intake of the parents. Tanner staging was not determined in the current study.

## CONCLUSION

The prevalence of obesity was higher in girls than in boys. The determinants of obesity include gender, age, source of income, SBP, LDL low HDL, Total trans fats and carbohydrates. Majority of overweight and obese adolescents led a sedentary lifestyle. The results observed in the study confirm the trend that is observed in urban children and adolescents. Lifestyle and diet modification should be encouraged to curb the onset of obesity. Nutrition and physical activity education components should be infused in the school curriculum to promote an active lifestyle. Department of Education together with health professionals and policymakers should develop a nutrition guide for food sold by vendors and revisit its school feeding scheme for optimal benefits. There is a need to conduct more studies to address gaps in government intervention programmes in South Africa with a view to promoting adolescent health.

## Data Availability

The data used for the research in this paper is owned by the University of Venda Research Ethics Committe.

## ACKNOWLEDGEMENTS

This work is based on research supported by the National Research Foundation (NRF) of SA, and the University of Venda, which are acknowledged for their contributions. The authors would also like to express their gratitude to the research team, the schools and learners who participated in the study

## AUTHOR CONTRIBUTION

**Conceptualization:** Brenda Baloyi and Solomon Mabapa.

**Data curation:** Brenda Baloyi

**Formal analysis:** Brenda Baloyi and Solomon Mabapa.

**Methodology:** Brenda Baloyi and Solomon Mabapa.

**Writing – original draft**: Brenda Baloyi and Solomon Mabapa

**Writing – review & editing:** Brenda Baloyi, Lindelani Fhumudzani Mushaphi and Solomon Mabapa.

## REFERENCES

1. World Health Organisation. Obesity. 2013. http://www.who.int/topic/obesity/en/.

2. Adom T, Puoane T, De Villiers A, Kengne AP. Prevalence of obesity and overweight in African learners: a protocol for systematic review and meta-analysis. BMJ open. 2017 Jan 1;7(1).

3. Gebrie A, Alebel A, Zegeye A, Tesfaye B and Ferede A. 2018. Prevalence and associated factors of overweight/obesity among children and adolescents in Ethiopia: a systematic review and meta-analysis. BioMed Central Journals, 5:19

4. Kimani-Murage EW, Kahn K, Pettifor JM, Tollman SM, Dunger DB, Gómez-Olivé XF and Norris SA. 2010. The prevalence of stunting, overweight and obesity, and metabolic disease risk in rural South African children. Bio Med central Public Health. 10:158. http://www.biomedcentral.com/1471-2458/10/158

5. Freedman DS, Mel Z, Srinivasan SR, Berenson and Dietz WH. 2007. Cardiovascular risk factors and excess adiposity among overweight children and adolescents: The Bagalusa Heart study. Journal of pediatric, 150: 12–17

6. World Health Organisation. 2018. Acting on childhood obesity. (World Obesity) Geneva, Switzerland

7. Niehues JR, Gonzales AI, Lemos RR, Bezerra PP, Haas P. Prevalence of overweight and obesity in children and adolescents from the age range of 2 to 19 years old in Brazil. International journal of pediatrics. 2014 Jun 3;2014.

8. Biadgilign S, Mgutshini T, Haile D, Gebremichael B, Moges Y, Tilahun K. Epidemiology of obesity and overweight in sub-Saharan Africa: a protocol for a systematic review and meta-analysis. BMJ open. 2017 Nov 1;7(11):e017666.

9. World health statistics. 2021: monitoring health for the SDGs, sustainable development goals. Geneva: World Health Organization; 2021. Licence: CC BY-NC-SA 3.0 IGO.

10. Chu DT, Nguyet NT, Dinh TC, Lien NV, Nguyen KH, Ngoc VT, Tao Y, Le DH, Nga VB, Jurgoński A, Tran QH. An update on physical health and economic consequences of overweight and obesity. Diabetes & Metabolic Syndrome: Clinical Research & Reviews. 2018 Nov 1;12(6):1095–100.

11. Alam MM, Hawlader MD, Wahab A, Hossain MD, Nishat SA, Zaman S, Ahsan GU. Determinants of overweight and obesity among urban school-going children and adolescents: a case-control study in Bangladesh. International journal of adolescent medicine and health. 2019 May 9;1(ahead-of-print).

12. World Health Organisation. 2021. https://www.who.int/news-room/fact-sheets/detail/obesity-and-overweight

13. Otitoola O, Oldewage-Theron W, Egal A. Prevalence of overweight and obesity among selected schoolchildren and adolescents in Cofimvaba, South Africa. South African Journal of Clinical Nutrition. 2020 Apr 8:1–6.

14. World Health Organisation. GLOBAL Strategy on Diet. Physical activity and Health. 2013b. http://www.who.int/dietphysicalactivity/childhood/en/.

15. World health organisation. The WHO Child Growth standards. 2013c. http://www.childgrowth/en/index.html.

16. Chooi YC, Ding C, Magkos F. The epidemiology of obesity. Metabolism. 2019 Mar 1;92:6–10.

17. Shisana O, Labadarios D, Rehle T, Simbayi L, Zuma K, Dhansay A, Reddy P, Parker W, Hoosain E, Naidoo P, Hongoro C, Mchiza Z, Steyn NP, Dwane N, Makoae M, Maluleke T, Ramlagan S, Zungu N, Evans MG, Jacobs L, Faber M, & SANHANES-1 Team. 2013. South African National Health and Nutrition Examination Survey (SANHANES-1). Cape Town: HSRC Press

18. Pedro TM, Kahn K, Pettifor JM, Tollman SM, and Norris SA. 2014. Under and overnutrition and evidence of metabolc disease risk in rural black South African children and adolescents. South African Journal of Clinical Nutritio.27(4):194–200

19. Muthuri SK, Francis CE, Wachira LJ, LeBlanc AG, Sampson M, Onywera VO, Tremblay MS. Evidence of an overweight/obesity transition among school-aged children and youth in Sub-Saharan Africa: a systematic review. PloS one. 2014 Mar 27;9(3):e92846.

20. Monyeki MA, Awotidebe A, Strydom GL, de Ridder JH, Mamabolo RL, and Kemper HCG. 2015. The challenges of underweight and overweight in South African children: Are we winning or losing the battle? A Systematic Review. International Journal of Environmental Research and Public Health. 12(1156-1173). Doi:10.3390/ijerph120201156

21. Malhotra R, Hoyo C, Østbye T, Hughes G, Schwartz D, Tsolekile L, Zulu J, Puoane T. Determinants of obesity in an urban township of South Africa. South African Journal of Clinical Nutrition. 2008 Jan 1;21(4):315–20.

22. Cole TJ and Lobstein T. 2012. Extended international (IOTF) body mass index cut-off for thinness, overweight, and obesity. Pediatric obesity. 7:284–294

23. Fernandez JR, Redden DT, Pietrobelli A and Allison DB. 2004. Waist circumference percentiles in nationally representative samples of African-American, European-American, and Mexican-American children and adolescents. Journal of paediatric. 145:439–344

24. World Health Organisation. 2011. Waist circumference and Waist-Hip Ratio. Report of a WHO expert consultation. 8-11 December 2008. Geneva, Switzerland. ISBN 9789241501491

25. Gibson RS. Principles of nutritional assessment. Oxford university press, USA; 2005.

26. Jolliffe CJ and Janssen, I. 2006. Distribution of Lipoprotein by age and gender in adolescents. Circulation Journal of American Heart Association, 114:1056–1062

27. The fourth report on the diagnosis, evaluation, and treatment of high blood pressure in children and adolescents. 2005. National Institute of Health. U.S. Department of Health and Human Services. NIH Publication No.05-5267

28. American Heart Association (AHA). 2005. New AHA Recommendations for Blood pressure measurement. American family physician. http://www.aafp.org/afp/2005/1001/p1398.html.

29. Sharkey BJ, Gaskill SE, 2007, Fitness and health 6th edition (Champaign, IL Human kinetics)

30. Debeila S, Modjadji P, Madiba S. High prevalence of overall overweight/obesity and abdominal obesity amongst adolescents: An emerging nutritional problem in rural high schools in Limpopo Province, South Africa. African Journal of Primary Health Care & Family Medicine. 2021;13(1).

31. Steyn, Nelia P., Johanna H. Nel, Sonia Malczyk, Linda Drummond, and Marjanne Senekal. “Provincial Dietary Intake Study (PDIS): Energy and Macronutrient Intakes of Children in a Representative/Random Sample of 1–< 10-Year-Old Children in Two Economically Active and Urbanized Provinces in South Africa.” International journal of environmental research and public health 17, no. 5 (2020): 1717.

32. UNICEF. Consultant for development of a national school nutrition guideline. Pretoria: UNICEF; 2018.

33. Negash S, Agyemang C, Metsha TE, Peer N, Erasmas RT, and Kengne AP. 2017. Differential prevalence and association of overweight and obesity by gender and population group among school learners in South Africa: A cross-sectional study. BioMedical Central Obesity, 4:29

34. Ford ND, Patel SA, and Narayan KM V. 2017. Obesity in Low- and Middle-Income Countries: Burden, Drivers, and Emerging Challenges. Annual Review of Public Health. 38:145–164. https://doi.org/10.1146/annurev-publhealth-031816-044604

35. Wells JC, Chomtho S, Fewtrell MS. Programming of body composition by early growth and nutrition. Proceedings of the Nutrition Society. 2007 Aug;66(3):423–34.

36. Wardle J, Haase AM, Steptoe A, Nillapun M, Jonwutiwes K, Bellisie F. Gender differences in food choice: the contribution of health beliefs and dieting. Annals of behavioral medicine. 2004 Apr;27(2):107–16.

37. Lenhart CM, Hanlon A, Kang Y, Daly BP, Brown MD, Patterson F. Gender disparity in structured physical activity and overall activity level in adolescence: evaluation of youth risk behavior surveillance data. International Scholarly Research Notices. 2012;2012.

38. Sekokotla MA, Goswami N, Sewani-Rusike CR, Iputo JE, Nkeh-Chungag BN. Prevalence of metabolic syndrome in adolescents living in Mthatha, South Africa. Therapeutics and clinical risk management. 2017;13:131.

39. Mogre V, Gaa PK, and Abukari RNS. 2013. Overweight, obesity and thinness and associated factors among school-aged children (5-14 years) in Tamale Northern Ghana. European Science Journal, 9(20). ISSN: 1857-7881

40. Mogre V, Nyaba R, Aleyira S, Sam NB. Demographic, dietary and physical activity predictors of general and abdominal obesity among university students: a crosssectional study. Springerplus. 2015 Dec;4(1):1–8.

41. Kelishadi R. Childhood overweight, obesity, and the metabolic syndrome in developing countries. Epidemiologic reviews. 2007 Jan 1;29(1):62–76.

42. Huxley R, Mendis S, Zheleznyakov E, Reddy S, Chan J. Body mass index, waist circumference and waist: hip ratio as predictors of cardiovascular risk—a review of the literature. European journal of clinical nutrition. 2010 Jan;64(1):16–22.

43. Micklesfield LK, Pedro TM, Kahn K, Kinsman J, Pettifor JM, Tollman S and Norris SA. 2014. Physical activity and sedentary behaviour among adolescents in rural South Africa: levels, patterns and correlates. BMC Public Health. 14(40). http://www.biomedcentral.com/1471-2458/14/40

44. WHO. 2016. Physical activity in adolescents. Fact Sheet. Europe

45. Biljon AV, McKune AJ, DuBose KN, Kolanisi U, and Semple SJ. 2018. Physical activity levels in urban-based South African learners: Across-sectional study of 7 348 participants. South African Medical Journal. 108(02): 126–131.DOI:10.7196/SAMJ.2018.v108i2.12766

46. Hanson SK, Munthali RJ, Micklesfield LK, Lobelo F, Cunningham SA, Hartman TJ, Norris SA and Stein AD. 2019. Longitudinal patterns of physical activity, sedentary behaviour and sleep in urban South African adolescents, Birth-To-Twenty Plus cohort. Hanson et al. BMC Paediatrics. 19(241). https://doi.org/10.1186/s12887-019-1619-z

47. Guthold R, Stevens GA, Riley LM, Bull FC. 2020. Global trends in insufficient physical activity among adolescents: a pooled analysis of 298 population-based surveys with 1·6 million participants. The Lancet Child and Adolescent Health. 4(1):23–35. https://doi.org/10.1016/S2352-4642(19)30323-2Get rights and content

48. World Health Organization, T., 2010. Global recommendations on physical activity for health. World Health Organization.

49. Lee IM, Shiroma EJ, Lobelo F, Puska P, Blair SN, Katzmarzyk PT, Lancet Physical Activity Series Working Group. Effect of physical inactivity on major non-communicable diseases worldwide: an analysis of burden of disease and life expectancy. The lancet. 2012 Jul 21;380(9838):219–29.

50. Kallio P, Pahkala K, Heinonen OJ, Tammelin TH, Pälve K, Hirvensalo M, Juonala M, Loo BM, Magnussen CG, Rovio S, Helajärvi H. Physical inactivity from youth to adulthood and adult cardiometabolic risk profile. Preventive Medicine. 2021 Apr 1;145:106433.

51. Tremblay MS, LeBlanc AG, Kho ME, Saunders TJ, Larouche R, Colley RC, Goldfield G, Gorber SC. Systematic review of sedentary behaviour and health indicators in school-aged children and youth. International journal of behavioral nutrition and physical activity. 2011 Dec;8(1):1–22.

52. Aryeetey R, Lartey A,Grace SM, Nti H, Colecraft E and Brown P. 2017. Prevalence and predictor of overweight and obesity among school-aged children in urban Ghana. BMC Obesity Journal. 4:38. DOI:10.118/s40608-017-0174-0

53. Ahmad A, Zulaily N, Shahril MR, Abdullah SEFH, and Ahmed A. 2018. Association between socioeconomic status and obesity among 12-year-old Malaysian adolescents. PLoS ONE 13(7): e0200577. https://doi.org/10.1371/journal.pone.0200577

54. Nirmal A, Venkataraman P, Kanniammal C, Rani A, Arulappan J. 2018. Prevalence of Obesity and Associated Risk Factors among Adolescents in Kancheepuram, South India. International Journal of Nursing Education, January-March. 10(1): 55–60.DOI 10.5958/0974-9357.2018.00012.0

55. Toriola AL, Moselakgomo VK, Shaw BS, and Goon DT. 2012. Overweight, obesity and underweight in rural black South African children. South African journal of clinical nutrition, 25(2):57–61

56. Kotain MS, Ganesh KS, and Kotian SS. 2010. Prevalence and determinants of overweight and obesity among adolescent school children of South Karnataka, India. Indian Journal of Community Medicine, 35(1): 176–178

57. Choukem SP, Kamdeu-Chedeu J, Leary SD, Mboue-Djieka Y, Nebongo DN, Akazong C, Mapoure YN, Hamilton-Sheild JP, Gautier J and Mbanya JC. 2017. Overweight and obesity in children aged 3-13 years in urban Cameroon: a cross-sectional study of prevalence and association with socio-economic status. BMC Obesity journal. 4:7. Doi 10.1186/s40608-017-0146-4

58. Kabbaoui EM, Chda A, Bousfiha A, Aarab L, Bencheikh R and Tazi A. 2018. Prevalence of and risk factors for overweight and obesity among adolescents in Morocco. East Mediterranean Health Journal. 24(6):512–521. https://doi.org/10.26719/2018.24.6.512

59. Roberts BP, Blinkhorn AS, Duxbury JT. The power of children over adults when obtaining sweet snacks. International Journal of Paediatric Dentistry. 2003 Mar;13(2):76–84.

60. van Ansem WJ, Schrijvers CT, Rodenburg G, van de Mheen D. Children’s snack consumption: role of parents, peers and child snack-purchasing behaviour. Results from the INPACT study. The European Journal of Public Health. 2015 Dec 1;25(6):1006–11.

61. Jung SH, Tsakos G, Sheiham A, Ryu JI, Watt RG. Socio-economic status and oral health-related behaviours in Korean adolescents. Social Science & Medicine. 2010 Jun 1;70(11):1780–8.

62. Ausems M, Mesters I, van Breukelen G, De Vries H. Do Dutch 11–12 years olds who never smoke, smoke experimentally or smoke regularly have different demographic backgrounds and perceptions of smoking?. The European Journal of Public Health. 2003 Jun 1;13(2):160–7.

63. Mohan S, Sarma PS, Thankappan KR. Access to pocket money and low educational performance predict tobacco use among adolescent boys in Kerala, India. Preventive medicine. 2005 Aug 1;41(2):685–92.

64. Chen CY, Lin IF, Huang SL, Tsai TI, Chen YY. Disposable income with tobacco smoking among young adolescents: a multilevel analysis. Journal of Adolescent Health. 2013 Jun 1;52(6):724–30.

65. Guo L, Xu Y, Deng J, He Y, Gao X, Li P, Wu H, Zhou J, Lu C. Non-medical use of prescription pain relievers among high school students in China: a multilevel analysis. BMJ open. 2015 Jul 1;5(7):e007569.

66. Ma J, Zhu J, Li N, He Y, Cai Y, Qiao Y, Redmon P, Wang Z. Cigarette smoking in Chinese adolescents: importance of controlling the amount of pocket money. Public Health. 2013 Jul 1;127(7):687–93.

67. Wang Y, Liang H, Tussing L, Braunschweig C, Caballero B, Flay B. Obesity and related risk factors among low socio-economic status minority students in Chicago. Public health nutrition. 2007 Sep;10(9):927–38.

68. Lachat C, Khanh LN, Khan NC, Dung NQ, Van Anh ND, Roberfroid D, Kolsteren P. Eating out of home in Vietnamese adolescents: socioeconomic factors and dietary associations. The American journal of clinical nutrition. 2009 Dec 1;90(6):1648–55.

69. Jensen JD, Bere E, De Bourdeaudhuij I, Jan N, Maes L, Manios Y, Martens MK, Molnar D, Moreno LA, Singh AS, te Velde S. Micro-level economic factors and incentives in Children’s energy balance related behaviours-findings from the ENERGY European cross-section questionnaire survey. International Journal of Behavioral Nutrition and Physical Activity. 2012 Dec;9(1):1–2.

70. Punitha VC, Amudhan A, Sivaprakasam P, Rathnaprabhu V. Pocket money: influence on body mass index and dental caries among urban adolescents. Journal of clinical and diagnostic research: JCDR. 2014 Dec;8(12):JC10.

71. Lee EY, Yoon KH. Epidemic obesity in children and adolescents: risk factors and prevention. Frontiers of medicine. 2018 Dec;12(6):658–66.

72. Ghomari-Boukhatem H, Bouchouicha A, Mekki K, Chenni K, Belhdj M and Bouchenak M. 2017. Blood pressure, dyslipidaemia and inflammatory factors are related to body mass index in scholar adolescents. Archives of Medical Science. 1:46–52. Doi:10.5114/aoms.2017.64713

73. Nkeh-Chungag B N, Sekokotla AM, Sewani-Rusike C, Namugowa A, Iputo JE. 2015. Prevalence of Hypertension and Pre-hypertension in 13-17-Year-Old Adolescents Living in Mthatha - South Africa: A Cross-Sectional Study. Central European Journal of Public Health. 23(1):59–64. DOI: 10.21101/cejph.a3922

74. Sebati RB, Monyeki KD, Monyeki MS, Motloutsi B, Toriola AL, and Monyeki MJL. 2019. Ellisras Longitudinal Study 2017: The relationship between waist circumference, waist-to-hip ratio, skinfolds and blood pressure among young adults in Ellisras, South Africa (ELS 14). Cardiovascular Journal of Africa.30(1): 24–28.DOI: 10.5830/CVJA-2018-056

75. Nguyen D, Kit B, and Carroll M. 2015. Abnormal Cholesterol Among Children and Adolescents in the United States, 2011–2014. NCHS Data Brief. Number 228

76. Bibiloni MM, Salas R, Novelo HI, Villarreal JZ, Sureda A, Tur JA. 2015. Serum Lipid Levels and Dyslipidaemia Prevalence among –10 Year-Old Northern Mexican Children. PLoS ONE 10(3):e0119877. doi:10.1371/journal.pone.0119877s

77. Miller WM, Nori-Janosz KE, Lillystone M, Yanez J, and McCullough PA. Obesity and Lipids. Current Cardiology Reports 2005. 7:465–470. ISSN 1523-3782

78. Daniels SR, Greer FR. 2008. Lipid Screening and Cardiovascular Health in Childhood. American Academy of Paediatrics. 122(1):198–208. DOI: 10.1542/peds.2008-1349

79. Wang H and Peng D. 2011. New insight into the mechanism of low high-density lipoprotein cholesterol in obesity. Lipids in health and disease. BioMed Central journal 10(176). http://www.lipidworld.com/content/10/1/176

80. Gupta R, Rao RS, Misra A and Sharma SK. 2017. Recent trends in epidemiology of dyslipidaemias in India. Indian Heart Journal. 69 (382-392). Doi.org/10.1016/j.ihj.2017.02.020

81. D’Adamo E, Guardamagna O, Chiarelli F, Bartuli A, Liccardo D, Ferrari F, and Nobili V. 2015. Atherogenic dyslipidemia and cardiovascular risk factors in obese children. International journal of endocrinology. 2015 (912047). https://doi.org/10.1155/2015/912047

82. Appannah G, Pot GK, Huang RC, Oddy WH, Beilin LJ, Mori TA, Jebb SA, Ambrosini GL. Identification of a dietary pattern associated with greater cardiometabolic risk in adolescence. Nutrition, Metabolism and Cardiovascular Diseases. 2015 Jul 1;25(7):643–50.

83. Allioua M, Djaziri R, Mahdad MY, Gaouar SB, Derradji H, Boudjemaa BM, and Belbraouet S. 2015. Dietary fat intake, micronutritient and obesity among adolescent in Tlemcen (Western Algeria). Food and Nutrition Sciences. 6(10):860.

84. Steyn N, Eksteen G, Senekal M. Assessment of the dietary intake of schoolchildren in South Africa: 15 years after the first national study. Nutrients. 2016 Aug;8(8):509.

85. Harris T, Malczyk S, Jaffer N, Steyn NP. 2019. How well are adolescents in the Gouda district of Western Cape meeting the South African food-based dietary guidelines for fat, sugar and sodium? Journal of Consumer Sciences. 4

86. Dreher ML. Role of fiber and healthy dietary patterns in body weight regulation and weight loss. Adv. Obes. Weight Manag. Control. 2015;3:00068.

87. Bahreynian M, Qorbani M, Motlagh ME, Riahi R, Kelishadi R. Association of dietary fiber intake with general and abdominal obesity in children and adolescents: The Weight disorder survey of the CASPIAN-IV Study. Mediterranean Journal of Nutrition and Metabolism. 2018 Jan 1;11(3):251–60.

88. Steyn NP, de Villiers A, Gwebushe N, Draper CE, Hill J, de Waal M, Dalais L, Abrahams Z, Lombard C, Lambert EV. Did HealthKick, a randomised controlled trial primary school nutrition intervention improve dietary quality of children in low-income settings in South Africa?. BMC Public Health. 2015 Dec;15(1):1–1.

